# Salivary molecular testing for SARS-CoV-2: simplifying the diagnosis without losing accuracy

**DOI:** 10.1101/2021.07.18.21260706

**Authors:** Francesca Saluzzo, Paola Mantegani, Valeria Poletti de Chaurand, Federica Cugnata, Patrizia Rovere-Querini, Marta Cilla, Patrizia Erba, Sara Racca, Cristina Tresoldi, Caterina Uberti-Foppa, Clelia Di Serio, Daniela Maria Cirillo

## Abstract

**Background:** Quantitative RT-PCR on NasoPharyngeal Swab (NPS) is still considered the standard for the diagnosis of SARS-CoV-2 infection, even if saliva has been evaluated in several studies as a possible alternative. The use of point of care (POC) platforms, providing highly specific results performed on saliva could simplify the diagnosis of COVID-19 and contribute to contain the spreading of SARS-CoV-2.

**Methods:** We assess the sensitivity and specificity of molecular testing performed on saliva in comparison to NPS using two different POC platforms (DiaSorin Simplexa™ and Cepheid Xpert®). NPS and saliva were collected prospectically from asymptomatic health care workers and mildly symptomatic patients. Moreover, the stability of saliva samples after storage at -80°C for up to 45 days was tested.

**Results:** The obtained results in comparison to NPS demonstrated for both DiaSorin Simplexa™ and Xpert® Xpress a specificity of 100% and a sensitivity of 90.24%. The overall agreement between the tests performed on saliva was 98%. A positive correlation in Ct values detected on saliva and on NPS was identified for all the targets shared by the tests in analysis (Orf1ab, E and N2). Both S Ct values and Orf1ab Ct values were not significantly different before and after the freezing in the tested saliva samples.

**Conclusion:** The obtained results demonstrated an overall performance of saliva comparable to NPS, confirming that RT-PCR performed using POCs on saliva could represent a valid public health solution for controlling SARS-CoV-2 pandemic.

## Introduction

The possibility to rely on rapid and accurate diagnostic techniques has proved itself crucial during the last year to contain the spreading of SARS-CoV-2 infection^1^. Even if quantitative RT-PCR (RT-qPCR) on NasoPharyngeal Swab (NPS) is still considered the standard for the diagnosis of COVID19, saliva has been evaluated in several studies as a possible alternative to NPS and it is now extensively utilized in South Korea, Germany, and Japan^2,3^. Nonetheless, the use of saliva is still debated, and a rigorous standardization of the analysis protocol is deeply needed ^4-6^. The application of point of care (POC) technologies on saliva, capable of rapidly performing high specific and sensitive molecular testing, could prove invaluable to allow the diagnosis even in challenging and remote settings, simplifying and speeding up the diagnostic process^1^.

## Methods

To assess the sensitivity and specificity of molecular testing performed on saliva in comparison to NPS using two different POC platforms (DiaSorin Simplexa™ and Cepheid Xpert®), a total 129 individuals were enrolled into the study. Samples were collected prospectically, from January 2021 to May 2021, from 21 asymptomatic health care workers, who took part to the screening campaign, and from 79 outpatients who had developed mild symptoms consistent with COVID19 up to 10 days before accessing the Preventive Medicine Unit, the COVID19 Mildly Symptomatic Outpatients Unit and the Emergency Department of San Raffaele Hospital, Milan. Moreover, samples from 29 patients, hospitalized for COVID19 in March 2020, were retrieved by San Raffaele Hospital Biobank. For each patient, a self-collected saliva sample and an NPS, collected at the same timepoint by a healthcare worker, were analysed.

With the exclusion of the samples collected in March 2020, that were stored at -80°C immediately after the sampling, all samples were preserved at 4°C and analysed within 24 hours from the collection. The DiaSorin Simplexa™ COVID-19 Direct tests (Simplexa™) were performed on saliva diluted 1:1 with saline as per the instruction for use, and the same condition was used off label for the Xpert® Xpress SARS-CoV-2 kit (Xpert®). NPS were analysed with Xpert® Xpress SARS-COV-2 or Roche Cobas® SARS-CoV-2 test (Cobas®), as by manufacturers’ instructions.

## Results

The results obtained on saliva samples collected prospectively in the first months of 2021, in comparison to NPS demonstrated for both Simplexa™ and Xpert® a specificity of 100% (95% CI: 93.94%-100%) and a sensitivity of 90.24% (95% CI: 76.87%-97.28%). The overall agreement between the two tests performed on saliva was of 98%.

Since the two kits employed on saliva, as well as the ones utilized on NPS, adopted different target genes (Simplexa™: Orf1ab and S; Xpert®: N2 and E; Cobas®: E and Orf1ab) an analysis to evaluate the correlation between Cycle threshold (Ct) values detected respectively on saliva and on NPS was performed only for the shared targets.

A positive correlation in Ct values detected on saliva and on NPS was identified for Orf1ab, detected both by Simplexa™ performed on saliva and by Cobas® on NPS (Kendall correlation = 0.7704, p <0.0001) as well as for E (Kendall correlation = 0.7961, p <0.0001) and N2 (Kendall correlation = 0.8311, p <0.0001), both targets of Xpert® performed on saliva and on NPS (Fig.1 A).

**Fig 1.**
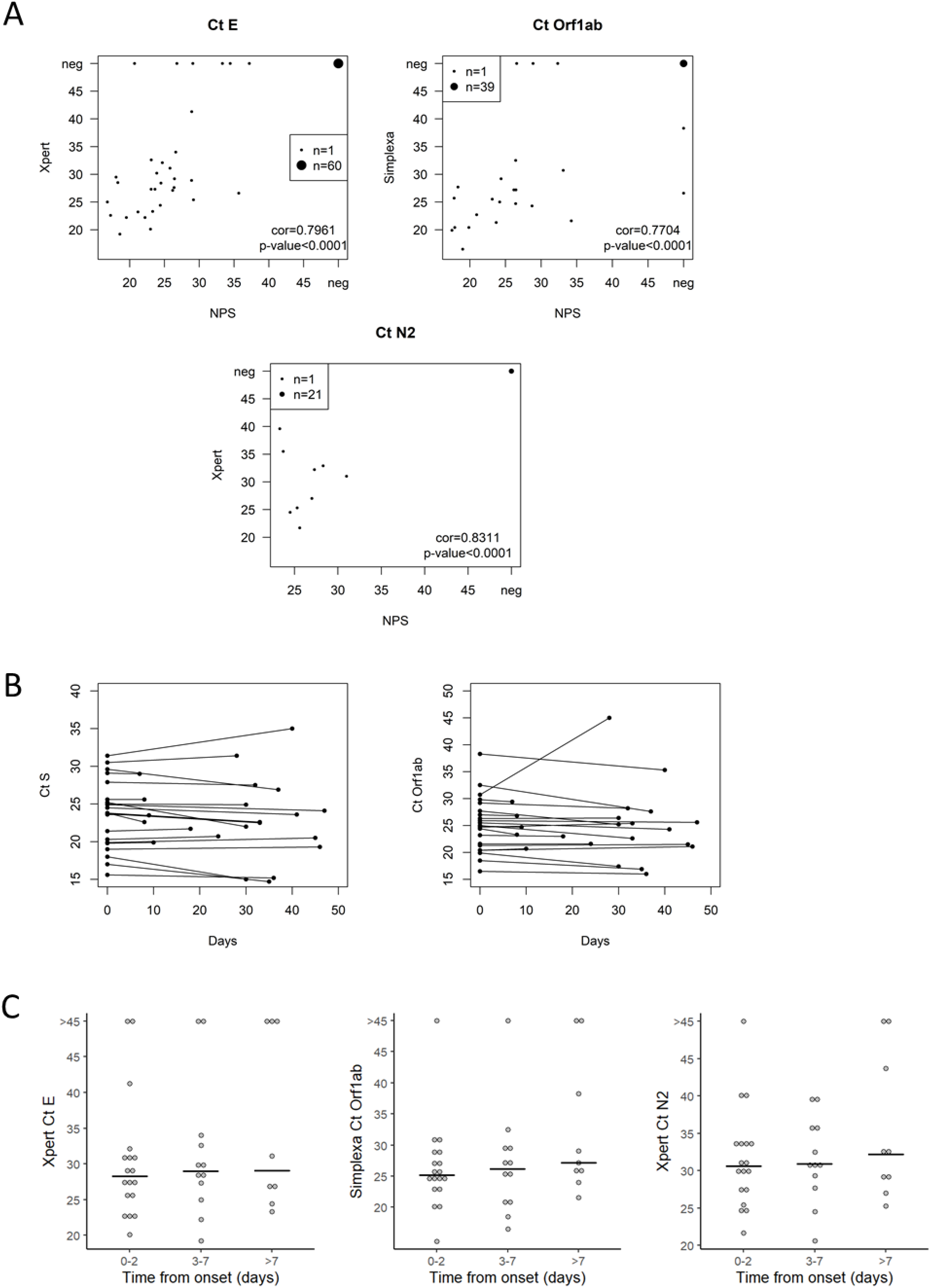
A) Ct values comparison between saliva (examined with Simplexa or Xpert) and NPS for different targets (genes S, Orf1ab and E); B) Ct values before and after storage at -80°C at different timepoints; C) Comparison of Ct values at different time frames from symptoms’ onset.

Hence, when compared to NPS, both tests on saliva freshly collected appeared to have good sensitivity and specificity as well as a positive correlation in Ct values detected for the shared targets.

Before proceeding to examine the samples collected in March 2020, we evaluated the effects of storage at -80°C for up to 45 days on 22 saliva samples that resulted positive for SARS-CoV-2. The samples were analysed with Simplexa™ before and after the freezing and the Ct values for the two different targets (S and Orf1ab) were compared. Both S Ct values and Orf1ab Ct values were not significantly different before (median (interquartile range): S Ct 23.8 (20.00–25.50), Orf1ab Ct 24.95 (21.38–27.52)) and after (median (interquartile range): S Ct 22.6 (20.55–25.4), Orf1ab Ct 24.5 (21.52–26.70) the freezing (Wilcoxon test 0.0705).

Moreover, the observed difference in Ct values did not appeared to be connected to the number of days for which the samples remained stored at -80 °C as a statistically significant correlation was not retrieved between the storage time and the Ct values (S: Kendall correlation p= 0.8206; Orf1ab: Kendall correlation p= 0.7128) (Fig. 1B).

Nonetheless, once the saliva samples collected by the Biobank from COVID-19 inpatients in March 2020 were included in the performance analysis (data not showed), the sensitivity resulted 87.14 (95% CI: 76.99-93.95) for Simplexa™ and 91.43 (95% CI: 82.27-96.79) for Xpert®, the agreement between Simplexa™ and Xpert® performed on saliva was 96.10%.

Considering that the median time from illness’ onset to collection of the Biobank specimens was 4 days (IQR: 2-9 days) while for the samples collected in 2021 was 2.5 days (IQR: 2-4 days), the time elapsed between symptoms’ onset and the samples collection was evaluated as a possible factor affecting Ct values and consequently cause the differences in specificity and sensitivity observed between the cohort of prospectically collected samples and the Biobank ones.

Categorizing the samples from symptomatic patients in three different categories (0-2 days from symptoms’ onset to collection, 3-7 days and >7 days) for saliva samples, no statistically significant differences were observed between different timeframes in Ct values for either of the target in analysis (E: Kruskal-Wallis test p=0.80; Orf1ab: Kruskal-Wallis test p=0.39; N2: Kruskal-Wallis test p=0.80) (Fig. 1C). Instead, NPS demonstrated an increase in Ct values, statistically significant for both E gene (Kruskal-Wallis test p= 0,007544) and for ORF1a/b gene (Kruskal-Wallis test p=0,03605).

## Conclusions

Both Xpert® and Simplexa™ platforms proved themselves practical and easy to use on saliva and the obtained results demonstrated an overall performance comparable to NPS, with a specificity of 100% and a sensitivity higher than 90% for freshly collected samples and higher than 87% for the ones stored at -80°C, demonstrating the possibility to perform these tests also on frozen samples with only a minimal loss in sensitivity. It is interesting to notice that the samples contained all the different SARS-CoV-2 variants of concern currently represented in Italy, but both kits’ performance was not compromised by this factor.

The tests employed exhibited an overall excellent level of agreement, even considering the differences identified once the Biobank samples have been included into the analysis.

As the pandemic evolves, the implementation of a testing strategy based on POCs widespread on the territory in a capillary manner, could help to guarantee a prompt on-site diagnosis, allowing the rapid identification and control of clusters and outbreaks, finally protecting the community from the transmission. Moreover, if this new diagnostic plan would involve the use of self-collecting highly reliable samples, as saliva, directly at patients’ homes, we would reduce the burden on healthcare workers, and the costs related to the use of NPS with specific transport medium. This approach would also contribute to drastically decreasing the number of possible infective individuals moving to reach the sampling hubs, who could represent a major public health risk.

This diagnostic approach could be easily implemented also in Low Middle Income Countries, where POCs platforms are already widely employed for the diagnosis of other illnesses, as Tuberculosis, HIV and viral hepatitis.

In conclusion, our findings support the use of saliva on POCs technologies as a valid solution to simplify, speed up and widespread the diagnostic process for the control of COVID19 epidemic.

## Data Availability

.

## Notes

### Competing Interest Statement

The authors have declared no competing interest.

### Funding Statement

.

### Author Declarations

IRCSS San Raffaele Ethics committee

